# Beyond T Staging in the Treat All Era: Capturing the Severity and Heterogeneity of Kaposi’s Sarcoma in East Africa

**DOI:** 10.1101/2020.01.04.20016519

**Authors:** Esther E. Freeman, Devon E. McMahon, Aggrey Semeere, Helen Byakwaga, Miriam Laker-Oketta, Megan Wenger, Charles Kasozi, Matthew Semakadde, Mwebesa Bwana, Michael Kanyesigye, Philippa Kadama-Makanga, Elyne Rotich, Job Kisuya, Kara Wools-Kaloustian, Ingrid Bassett, Naftali Busakhala, Jeffrey Martin

**Affiliations:** Massachusetts General Hospital, Harvard Medical School, Boston, Massachusetts, USA; Infectious Diseases Institute, Makerere University, Kampala, Uganda; University of California, San Francisco, USA; Masaka Regional Referral Hospital, Uganda; Mbarara Regional Referral Hospital, Uganda; Acaedmic Model Providing Access to Healthcare (AMPATH), Eldoret, Kenya; Indiana University, Indianapolis, Indiana, USA; Moi University, Eldoret, Kenya

## Abstract

**Background:** In the treat-all era of HIV, Kaposi’s sarcoma (KS) remains one of the most incident cancers in sub-Saharan Africa. The majority of patients with KS are diagnosed at advanced disease stage in this setting. Staging systems for KS, specifically the AIDS Clinical Trials Group (ACTG) system, were developed in the pre-ART era, were not meant to guide treatment, and may not fully capture the clinical heterogeneity of advanced disease. There is no international consensus on which KS patients need chemotherapy in addition to antiretroviral therapy (ART). Understanding KS severity of disease in the current era would help to inform prognosis and clarify treatment guidelines.

**Methods:** We performed rapid case ascertainment (RCA) on people living with HIV ≥18 years old newly diagnosed with biopsy-proven KS from 2016 to 2019 at three clinic sites in Kenya and Uganda. As close as possible to time of diagnosis, we performed a structured interview, physical examination, and collection of laboratory specimens. We reported KS severity using ACTG and WHO staging criteria, as well as detailed measurements not captured in current staging systems.

**Results:** We enrolled 264 adults newly diagnosed with KS. RCA was performed within 1 month of KS diagnosis for 62% of patients and within 6 months for 73% of patients. Patients were 61% Kenyan, 69% male, and with a median age of 35. Median CD4 count was 239 (IQR 87 to 408), with 72% of patients initiating ART greater than 60 days prior to diagnosis. The majority of patients had advanced stage of disease, with 82% qualifying as ACTG T1 and 64% as WHO Severe/Symptomatic KS. There was marked heterogeneity within advanced KS, with 25% of patients having two ACTG qualifiers and 3% of patients had three or more ACTG qualifiers.

**Conclusion:** The majority of patients with KS in this study had advanced stage disease at time of diagnosis, highlighting the need to improve early diagnosis of KS. Within this group of advanced stage patients was large clinical heterogeneity, leading to questions about whether all patients with advanced KS require the same treatment strategy.

## Introduction

Despite increasing availability and use of antiretroviral therapy (ART) in the treat all era for HIV, Kaposi’s sarcoma (KS) continues to be the most incident cancer in many countries in eastern and southern Africa.^1,2^ In these settings, one year survival for patients with KS is 65-79%,^3-5^ with 69-92% of patients presenting with T1 disease^3,4,6-8^ Staging for KS in sub-Saharan Africa is based on the AIDS Clinical Trials Group (ACTG) system, which American oncologists developed in 1989 to facilitate clinical trials for KS in high-income settings.^9,10^ ACTG staging criteria classifies patients into good- or poor-risk groups based on tumor extent (T), immune system status (I) measured by CD4+ count, and systemic illness (S) measured by evidence of HIV-related illness (Figure 1).^9^ Since the introduction of highly active ART in 1995, a number of significant changes in the treatment and epidemiology of KS have led to questions about whether ACTG staging is still optimal.^3,11^ To address these limitations, in 2014 the WHO developed a modified staging system for KS specifically to guide treatment in resource limited settings, which is not yet in wide use, and has yet to be validated or compared to ACTG staging.^12^

**Figure 1:**
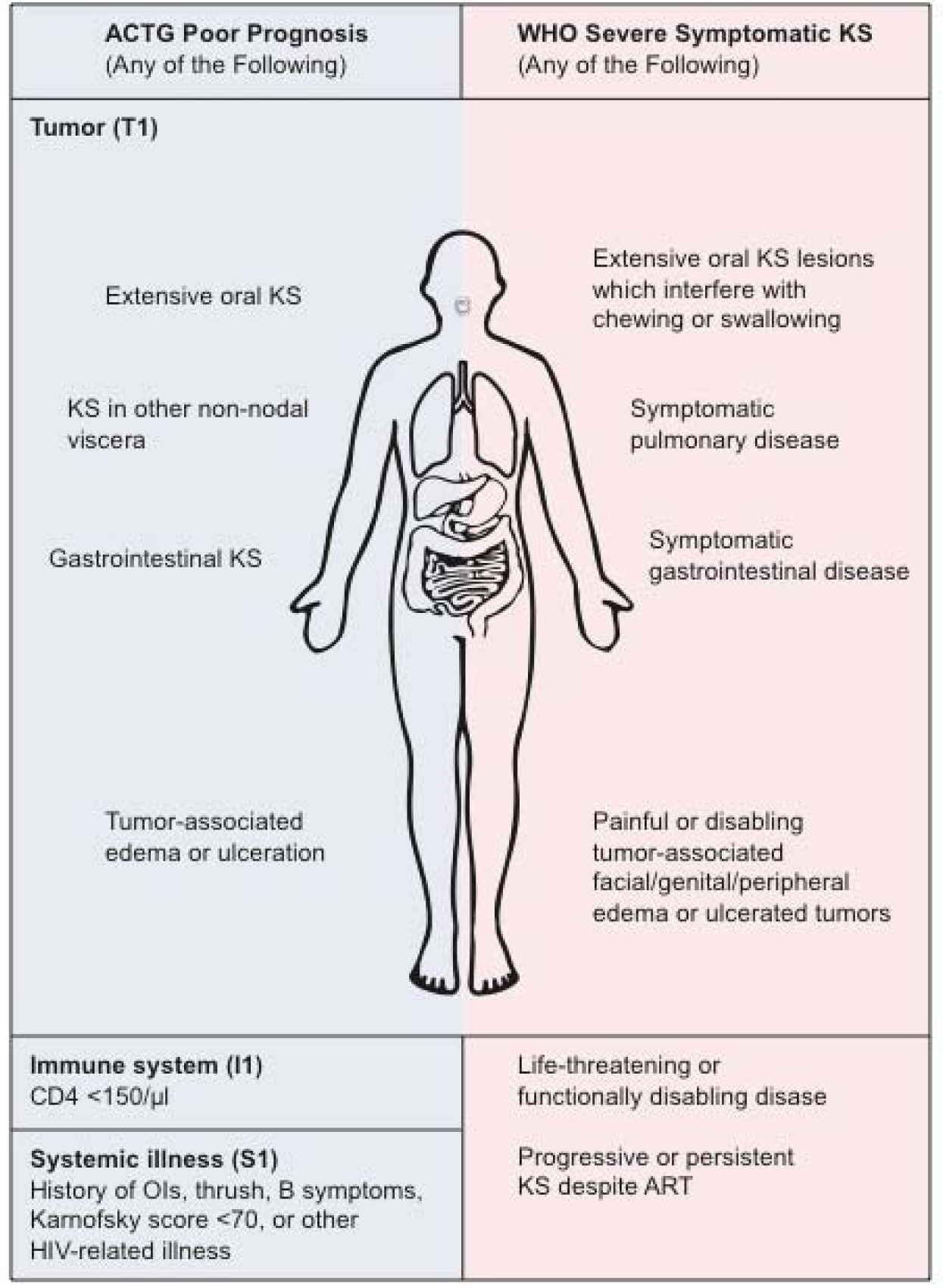
KS staging – a comparison of AIDS Clinical Trials Group (ACTG) and WHO KS Treatment Guidelines staging.

Ultimately, a cancer staging system should be both of prognostic value and guide therapy. Currently, there is no universally accepted standard for which patients with KS should receive chemotherapy.^13^ In resource rich settings, the National Comprehensive Cancer Network (NCCN) guidelines recommend using ACTG staging to dictate that T0 patients should receive ART only, and T1 patients should receive ART plus liposomal anthracycline chemotherapy.^14,15^ This guidance may work less well in resource limited settings, where most patients have advanced disease^4^ and liposomal anthracyclines are not readily available.^16^ Given the large burden of T1 disease, there are likely gradations of tumor burden that are not currently being captured in the ACTG staging system. Parsing out these potential gradations within T staging in sub-Saharan Africa has yet to be performed, and will likely have important implications for therapy. For example, there may be patients with T0 disease who would benefit from earlier intervention with chemotherapy, while there may be patients with T1 disease who would have disease remission on ART alone. To better understand this variation, more thorough measurements of KS at time of diagnosis, particularly for the large burden of advanced KS, is needed.

Using a rapid case ascertainment approach and detailed physical examination, biologic measurements, and self-reported symptoms, we assessed KS severity as close as possible to the time of KS diagnosis in a real-world, community setting in Kenya and Uganda. Better characterizing stage at diagnosis is important for further understanding KS diagnostic delays, prognosis, and appropriate treatments.

## Methods

### Overall Design

We used a rapid case ascertainment (RCA) approach at HIV primary care clinics in Kenya and Uganda to identify all patients newly diagnosed with KS from 2016 to 2019. RCA is designed to identify cases as soon as possible after the patient develops a disease, to allow for real-time investigation of cases with rapidly changing disease states. RCA is often used in high income settings to evaluate rapidly progressive cancers, and has been used previously in Africa to document infectious diseases outbreaks.^17-19^ We ascertained potential new cases of KS through the following mechanisms: i) regular queries of the electronic medical record for KS diagnosis or symptoms possibly attributable to KS, ii) regular scanning of pathology reports at the pathology department, iii) regular reviews of oncology patient registers, iv) regular reviews of dermatology clinic registers, and iv) clinician notification via telephone hotline. Upon identification of a patient with possible new KS diagnosis, the study team then contacted the patient either in-person or by telephone to determine eligibility and plan an RCA encounter. This approach allowed us to quickly evaluate patient characteristics, the extent of KS, and its complications as close as possible to the moment of initial KS diagnosis.

### Study Population

Inclusion criteria were people living with HIV (≥18 years old) receiving care at one of three HIV primary care networks in Kenya and Uganda, who were newly diagnosed with pathologically confirmed KS. Enrollment began in June 2016 in Western Kenya through the Academic Model Providing Access to Healthcare (AMPATH) clinic network, which in 2016 had 160,000 people living with HIV receiving care at over 30 sites. We then added two additional sites in January 2018 in Uganda: The Immune Suppression Syndrome (ISS) Clinic in Mbarara and the AIDS Healthcare Foundation (AHF) Uganda Cares Clinic in Masaka. At the time of study initiation, the ISS Clinic in southwestern Uganda had 10,889 active adult patients with HIV, while AHF Uganda Cares in southcentral Uganda had 13,317 patients with HIV. All sites provide ART free at charge, and there are affiliated oncology and dermatology clinics at AMPATH and ISS Clinic.

All three sites are part of the East Africa International Epidemiologic Databases to Evaluate AIDS (IeDEA) Consortium, which has electronic databases to document ambulatory HIV clinic data and provided skin biopsies free of charge for KS diagnosis. All patients provided written consent for data derived from their care at the participating clinic sites to be used for purposes of research through International Epidemiology Databases to Evaluate AIDS (IeDEA). Approval for this research was granted by the Institutional Research Ethics Committee (IREC) at Moi University in Eldoret, Kenya (Approval No.0001827) and by the Makerere University College of Health Sciences School of Biomedical Sciences Higher Degrees Research and Ethics Committee (SBS-HD-REC-495) in Uganda, as well as the University of California, San Francisco. Secondary data analysis was approved by Partners Human Research Committee (the Institutional Review Board of Partners Healthcare, Boston, Massachusetts).

### Measurements

#### Kaposi’s Sarcoma

Diagnosis of KS was made with punch biopsy and histologic confirmation, using anti-Lana staining. Kenyan and Ugandan pathologists with specialty training in dermatopathology performed an initial read. Dermatopathologists at UCSF performed a second, confirmatory read. After patients were determined eligible for RCA, clinical officers trained as research assistants performed a structured interview, physical exam, and collected biologic samples for laboratory evaluation. Questionnaires were used to evaluate potential signs or symptoms of cutaneous, oral, gastrointestinal, and pulmonary KS, as well as the patient’s experience of these symptoms and the impact of the KS symptoms on their activities of daily life. Similarly, the location and severity of edema, as well as the patient’s experience of their edema and its impact on their activities, were assessed. For each symptom category, the research assistant made an overall clinical determination whether reported symptoms were “definitely (>95%),” “probably (60-94%),” “possibly (30-59%),” “probably not (5-29%)” or “not (<5%)” related to KS. Additionally, the research assistant performed an overall Karnofsky performance scale (KPS) measurement. All patients were then fully examined, including a full skin and lymph node exam with removal of clothing. For suspected KS lesions, measurements were made using a standard body diagram of cutaneous and oral involvement, lesion counts, lesion dimensions, and presence of ulceration or infection. Location and dimensions of edema were also recorded. In addition, detailed photographs were taken with patient consent. Tumors were staged according to ACTG staging.^9^ KS was also staged according to WHO KS Treatment Guidelines staging. ^20^

#### Other Laboratory Measurements

Blood samples for laboratory collection were collected to determine hemoglobin, HIV viral load, and CD4+ T cell count.

### Statistical Analysis

Patient characteristics were evaluated at time of RCA enrollment, which was as soon as possible after confirmation of diagnosis. For the subset of patients newly diagnosed with KS while already on ART, we evaluated whether they were virally suppressed at enrollment (HIV RNA <200 copies/ml). For binary variables, we estimated proportions and confidence intervals. For continuous variables, we summarized median and interquartile range (with confidence intervals). All analyses were performed using Stata (version 16.0, Stata Corp., College Station, Texas).

## Results

### Characteristics of the Study Population

We performed rapid case ascertainment on 264 people living with HIV with newly diagnosed KS from 2016 to 2019. Their median age was 35 and males comprised 69% of the sample (Table 1). Patients were enrolled from AMPATH-Kenya (69%) as well as ISS-Uganda and AHF Uganda Cares-Uganda (31%). RCA was performed within 1 month of KS diagnosis for 62% of patients and within 6 months for 73% of patients, with the main reason for inability to perform RCA being intervening death (66%). Median Karnofsky Performance Scale score was 70 (IQR 60 to 80) and Hemoglobin was 11 g/dl (IQR 9.1 to 12.8). Patients presented with a median CD4 count of 239 (IQR 87 to 408) with 90% of patients on ART at time of study enrollment and 72% of patients using ART greater than 60 days prior to diagnosis. The majority of patients had advanced stage of disease, with 82% qualifying as ACTG T1 and 64% as WHO Severe/Symptomatic KS.

**Table 1.** Characteristics of people living with HIV newly diagnosed with KS in Kenya and Uganda at the time of study enrollment.

| **Characteristic** | **Median (IQR) or Percentage**<br>**(N=264)** |
| --- | --- |
| **Age** | 35 (30 to 42) |
| **Male sex** | 31% |
| **Education level** |  |
| None/Primary | 66% |
| Secondary | 28% |
| Tertiary/University | 6% |
| **Literate\*** | 83% |
| **Country** |  |
| Kenya | 61% |
| Uganda | 39% |
| **Stage** | 120 (50 to 270) |
| ACTG T1 | 82% |
| WHO Severe KS | 64% |
| **Weight** |  |
| **Karnofsky Performance Scale** | 70 (60 to 80) |
| **ART status** |  |
| On ART at time of staging | 90% |
| On ART >60 days prior to diagnosis | 72% |
| **Hemoglobin, g/dl** | 11 (9.1 to 12.8) |
| **CD4+ T-cells, cells/mm3** | 239 (87 to 408) |
| CD4 <150 | 65% |
| **Viral load categories, copies/ml** |  |
| undetectable (<40) | 48% |
| 40 – 1,000 | 35% |
| 1,001 – 10,000 | 11% |
| >10,000 | 6% |

\* Participants were considered literate if they were able to read three sentences of Grade-5 level writing in their primary language.

### Reported Symptoms at Time of Diagnosis

Many patients reported constitutional symptoms, including fever (34%), chills or sweats (31%), diarrhea (13%), loss of appetite (41%), and weight loss (34%). Patient-reported oral symptoms included difficulties with swallowing, eating, drinking or speaking (17%), bleeding in the mouth (9%), and pain in the mouth (13%) (Table 2). Patient-reported gastrointestinal symptoms comprised abdominal pain (21%), vomiting (11%), throwing up blood (1%), blood in the stool (7%), and feeling full prematurely (27%). Patient-reported pulmonary symptoms included shortness of breath (22%), cough (28%), and coughing up blood (4%). Patients additionally reported extremity edema (77%), periorbital edema (12%), and genital or scrotal edema (22%).

**Table 2.**
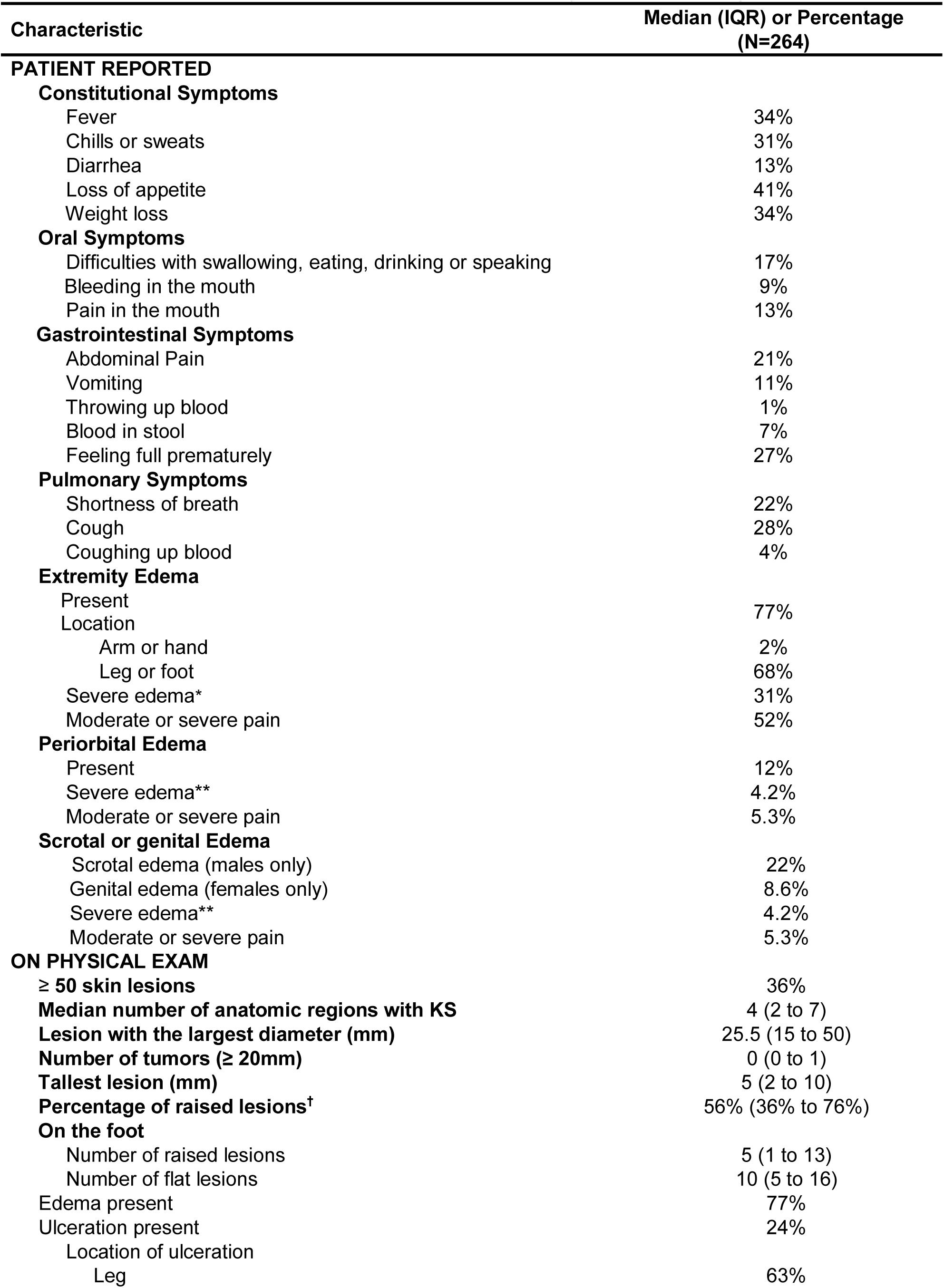
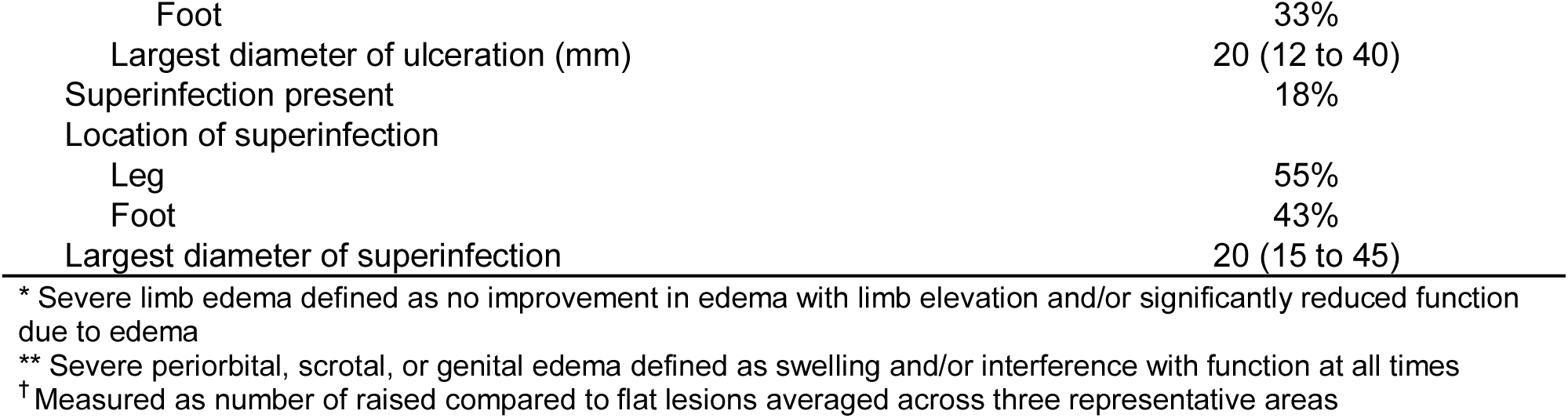
Patient reported and clinician documented symptoms and signs related to KS

### Exam Documentation at Time of Diagnosis

On clinician-documented physical exam, all patients had cutaneous lesions, with 36% demonstrating ≥ 50 lesions. The largest diameter of lesions was 25.5 mm (IQR 15 to 50), with the tallest lesions measuring 5 mm (IQR 2 to 10). When comparing the number of raised to flat lesions averaged across three representative areas of KS, the median percentage of raised lesions was 56% (IQR 36% to 76%). The most common sites of cutaneous KS included the legs (76%), feet (64%), arms (48%), back (39%), head (38%), chest (35%), oral cavity (34%) abdomen (32%), hands (30%), neck (29%), genitals (28%), and buttocks (24%) (Figure 2). Of the 34% of patients with oral KS, the most common sites included the hard palate (81%), soft palate (50%), buccal mucosa (34%), gingiva/gums (29%), tongue (22%), floor of the mouth (10%), posterior pharynx (5%), tonsils (4%) and uvula (4%). Ulceration was documented in 24% of lesions and superinfection was documented in 18% of lesions. Edema was presents in 77% of patients, with the most common locations including the feet (77%), legs (73%), genitals (14%), face including eyes (7%), hands (4%) and arms (3%). For patients with lower extremity edema, median size measured across the foot was 26.2 cm (IQR 24.3 to 28.8), across the lower leg was 28.4 cm (IQR 24.4 to 34.0) and across the thigh was 46.0 cm (IQR 40.0 to 50.7 cm).

**Figure 2:**
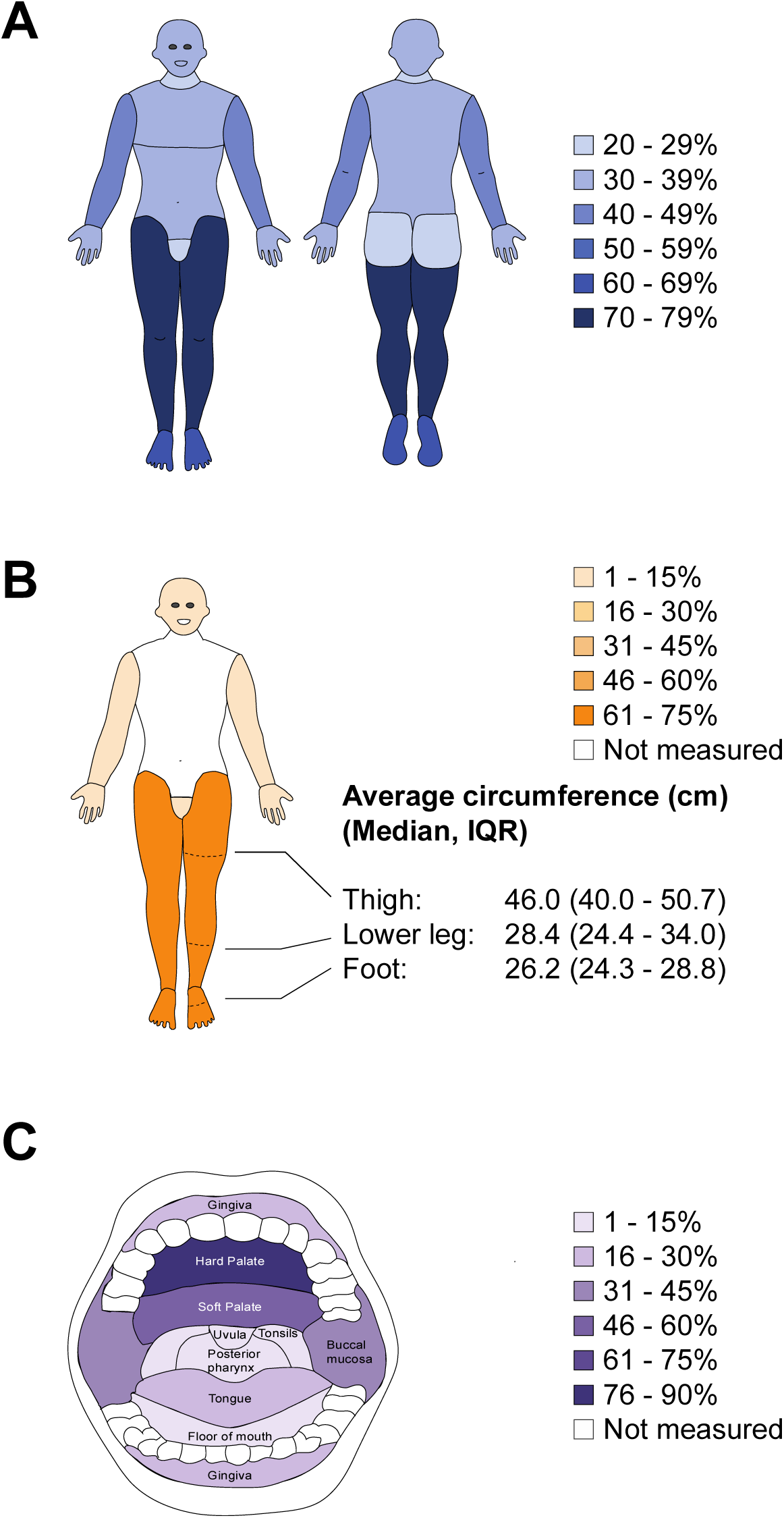
Distribution of KS-related A) Cutaneous Lesions, B) Edema, and C) Oral Lesions at enrollment, as close as possible to moment of KS diagnosis.

### KS Staging at Time of Diagnosis

For ACTG staging, 18% of patients were ACTG T0 and 82% ACTG T1. 54% of patients had one qualifier for ACTG T1, while 25% of patients had two qualifiers and 3% of patients had three or more qualifiers (Figure 3). The most common qualifying symptoms for ACTG T1 were tumor associated edema (70%), tumor associated ulceration (24%), extensive oral KS (9%), pulmonary KS (7%), and gastrointestinal KS (4%) (Figure 4) For patients with two or more ACTG T1 qualifiers, the most common symptom combinations included edema plus ulceration (n=54) and edema plus oral KS (n=12). For WHO staging, 36% of patients had WHO Mild/Moderate KS while 64% of patients had WHO Severe KS. 42% of patients had one qualifier for WHO Severe KS, 15% had two qualifiers, and 7% had three or more qualifiers. The most common qualifying symptoms for WHO Severe were painful or disabling edema or ulcerated tumors (46%), life threatening or functionally disabling disease (22%), extensive oral KS interfering with chewing or swallowing (13%), symptomatic gastrointestinal disease (8%) and symptomatic pulmonary disease (7%). For those with two or more WHO Severe KS, the most common symptom combination was edema plus life-threatening/disabling symptoms (n=34).

**Figure 3:**
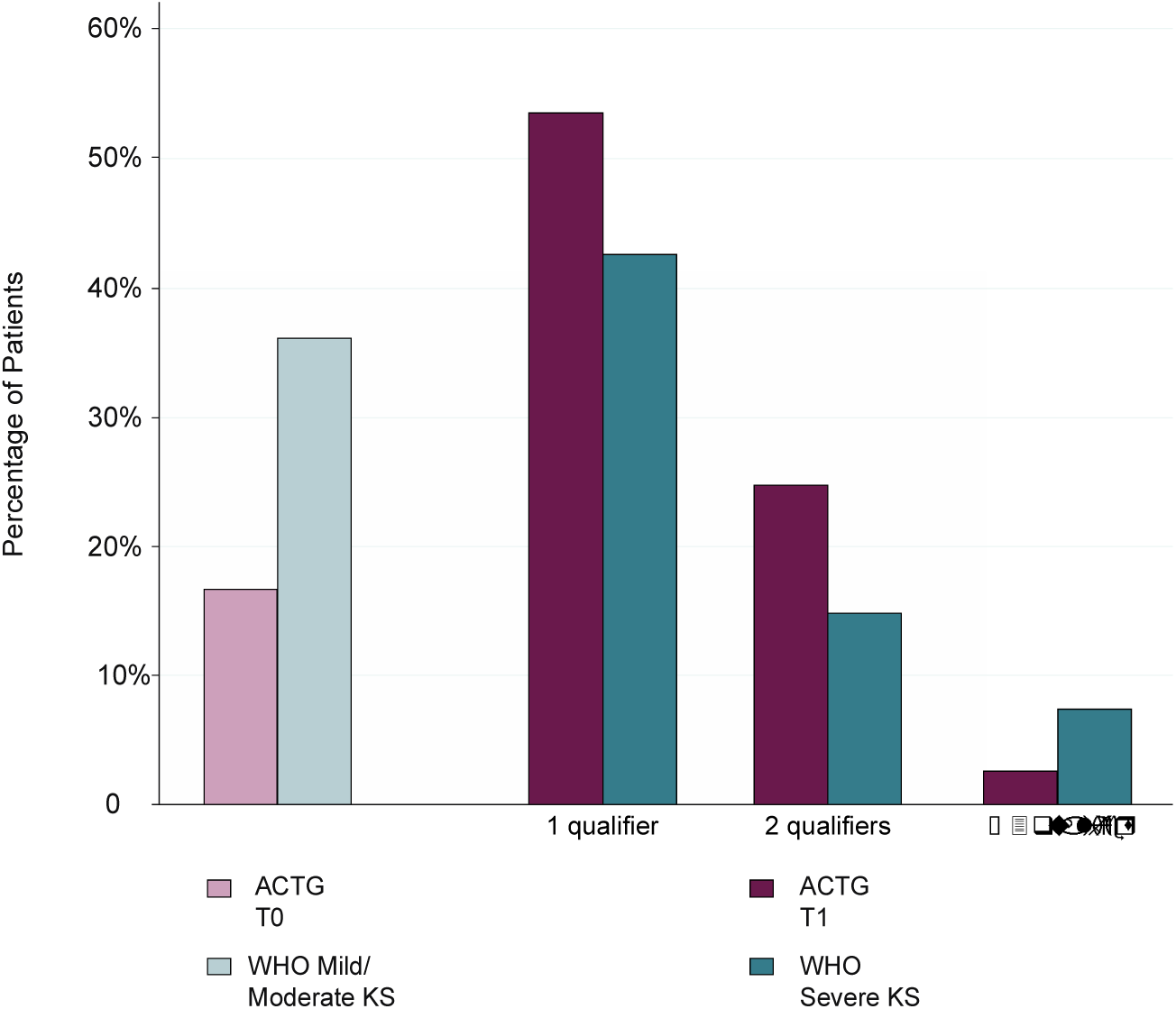
Proportion of patients with symptoms qualifying them for ACTG T1 KS or WHO “severe” KS.

**Figure 4:**
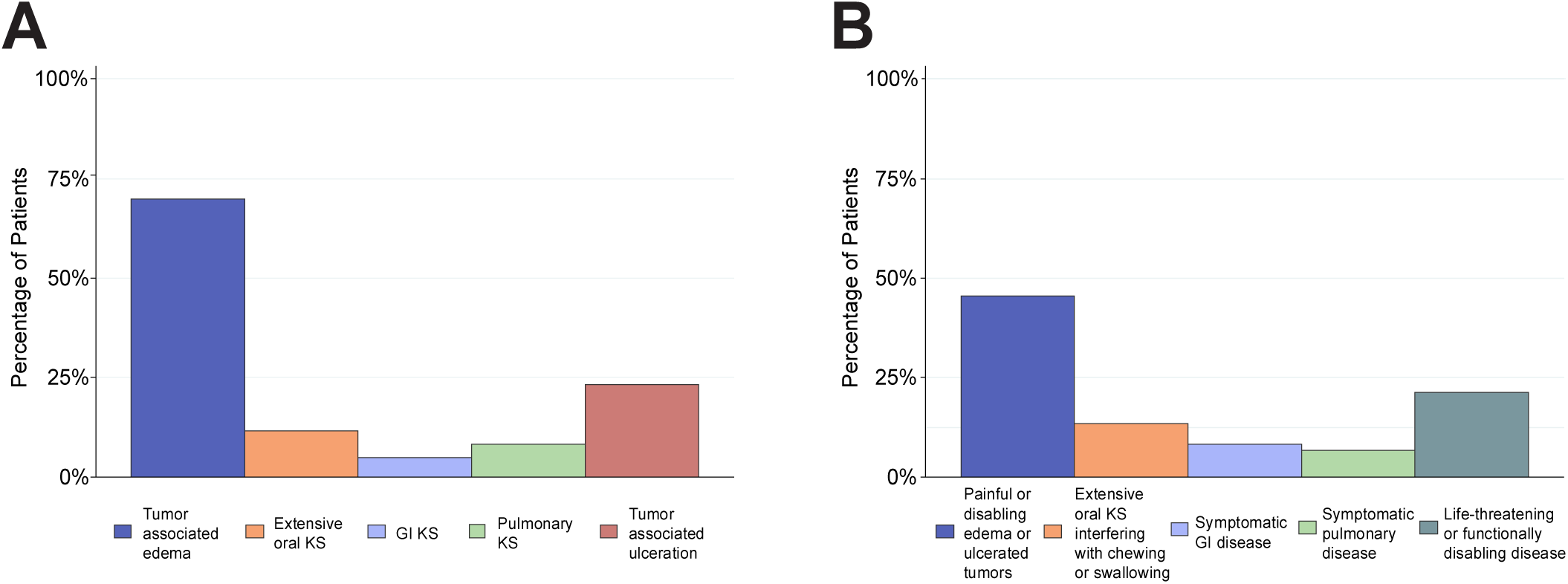
Symptoms qualifying patients for A) ACTG T1 KS or B) WHO “severe” KS.

## Discussion

This study provides a comprehensive assessment of disease stage among a prospective community-based sample of HIV-infected adults newly diagnosed with KS in East Africa. Rapid case ascertainment was used to obtain detailed measurements on 62% of people living with HIV within 1 month of first KS suspicion, making this study to our knowledge the most thorough and proximal assessment of KS stage at diagnosis in sub-Saharan Africa. By using detailed patient interviews, physical examinations, and laboratory evaluations, we were able to demonstrate substantial clinical heterogeneity among patients with advanced KS, particularly within ACTG T1 stage of disease.

The majority of patients in our study were diagnosed with advanced disease, with 82% of patients having ACTG T1 disease and 64% having WHO Severe/Symptomatic KS. These findings are similar to prior studies in sub-Saharan Africa reporting that 69 to 89% of KS patients present with T1 disease.^4,5,7,21,22^ This finding contrasts with high income countries where there is much less T1 disease, with recent estimates of 34-35%.^14,23^ Compared to prior studies, our documentation of advanced stage of disease is likely more representative and proximal in a multi-site sub-Saharan African setting. In contrast, the majority of prior studies on KS stage in sub-Saharan Africa relied on chart review data, which often has missing records and incomplete documentation to determine staging.^3,5,7,21,22^ Other studies used clinical trial data or prospective cohort data to report stage, which contained rigid inclusion criteria and often excluded the sickest patients.^4,6,8^

Another highlight of this study was our ability to document the clinical variability hidden within the 82% of patients with ACTG T1 stage, which until now has not been reported in the literature. The most common qualifying symptom for ACTG T1 was tumor associated edema in 70% of patients, though notably 25% of patients had two ACTG qualifiers and 3% of patients had three or more qualifiers. Similarly, for WHO Severe staging 15% of patients had two qualifiers and 7% of patients had three or more qualifiers. Additionally, a smaller percentage of patients qualified for WHO Severe/Symptomatic KS compared to ACTG T1 KS, an effect mostly mediated by the classification of tumor-associated edema. While 70% of patients qualified for ACTG T1 due to “tumor associated edema,”, only 46% qualified for WHO Severe/Symptomatic KS due to “painful or disabling edema or ulcerated tumors.”

The WHO staging criteria focuses on the impact of KS symptoms on function and quality of life, while ACTG staging focuses on the presence or absence of KS symptoms. The differences we found between ACTG and WHO staging are important, as the diagnosis of advanced KS currently delineates which patients are prescribed chemotherapy. Classifying tumor burden in KS is challenging as there is no “primary” site.^24^ These uncertainties about staging have direct implications for KS treatment, as there continues to be ambiguity regarding which subtype of patients would benefit from chemotherapy in addition to ART. For example, in one study from South Africa although only 15% of KS patients were T0, 39% of patients responded to ART alone.^4^ Given the immense heterogeneity described among T1 patients in our study, we remain skeptical that all patients classified as “T1” require the same treatment approach. For example, some patients with T0 KS may have extensive skin disease requiring chemotherapy, and some patients with T1 KS may have mild edema not requiring chemotherapy. Especially in resource limited areas with higher rates of opportunistic infection, loss to follow-up, and decreased capacity for supportive care, both under treatment with ART alone and overtreatment with chemotherapy are potentially perilous.

The reason for the large burden of T1 KS in sub-Saharan Africa is at least partially due to diagnostic delays, both during the time interval between patients noticing symptoms and reaching a clinician (“primary delay”), as well as between seeing a clinician and obtaining a diagnosis (“secondary delay”).^8^ A study from Uganda showed that 45% of KS patients had a primary delay of greater than 3 months due to lack of pain, lack of money for transportation, and far distance to the facility. Patients who had a primary delay of three or more months were three times as likely to present with ACTG poor risk stage.^8^ This study further found that KS patients already enrolled in a HIV primary care clinic had the same diagnostic delay interval as those not accessing care, suggesting a lack of screening and knowledge of KS among HIV workers.^8^ This underscores the need to improve knowledge of HIV-associated malignancies among frontline HIV cadres, as well as incorporate cancer screening with routine HIV services. Especially in settings with limited ability to provide first line chemotherapy, early diagnosis of KS is critical.

The proximity of our assessment of stage to time of diagnosis additionally provided unique insights about the natural course of KS. The lower limbs were the most common sites of cutaneous KS.^5,7^ Similar to one prior study we found that only 34% of patients presented with oral KS^5^, which is lower than the 58%-65% reported in other studies.^7,25^ Comparable to prior studies we found a large number of lesions that were either ulcerated or superinfected.^3^ We demonstrated that 77% of patients presented with edema with the most common location being the lower limbs and genitals, which has also been consistently reported in prior literature.^3,25^ We demonstrated that 7% of patients presented with pulmonary KS and 8% presented with GI KS, which is similar to reports from prior retrospectives studies.^5,7^

Designing a staging system for HIV-associated KS presents unique challenges in the era of ART, where many patients are already on ART at time of diagnosis. In this cohort, 72% of patients were on ART greater than 60 days prior to KS diagnosis and 48% had an undetectable viral load, underscoring the burden of KS despite ART. We surmise that the magnitude of disease burden has heightened importance in this setting, and that immune status (I) and systemic symptoms (S), which are greatly modified by ART, ^3,4,8,19^ may be less important now than they were during the development of the ACTG staging system in the 1980s.

However, the question remains which signs or symptoms of KS are predictive of KS mortality. In addition to TIS staging predictors, previous studies from both high and low income settings have found that pulmonary involvement,^11^ woody edema,^22^ low BMI,^26^ low hemoglobin,^26^ low albumin,^25^ older age,^23^ decreased performance status,^23,25^ and positive HHV-8 DNA to be predictive of mortality.^6,27^ The linkage of staging criteria with outcomes has recently been performed in the pediatric KS literature in Malawi, but has not yet been performed for the adult KS population in sub-Saharan Africa during the treat-all era.^28^ Our next steps will be to link the detailed measurements from this study on stage of disease in the ART era with mortality of these patients.

Our study has several limitations. The majority of eligible patients were enrolled within 1 month of diagnosis (62%), however some patients were enrolled up to six months after diagnosis, and some eligible patients were not captured. The main reason for not capturing patients was intervening death (66%), thus our study likely under represents disease burden at time of diagnosis. Additionally, there were unique facilitators at our sites that may not be generalizable to other locations. All of the sites were part of the IeDEA network, which provided KS biopsy services free of charge, allowing for a more rapid diagnosis. Additionally, all participating sites were part of a HIV-primary care network, thus a larger percentage of our patients may have been on ART compared to other areas. Because this study was conducted across multiple sites, there is likely some variability in how measurements were taken. Furthermore, this study relied on both self-reported measures as well as physical exam-documented measures, which were not always concordant. This was especially problematic for measuring visceral KS involvement, as we were unable to endoscopically confirm GI or pulmonary KS. Chest X-Rays, CT scans, and endoscopies were not routinely collected in our patients as part of standard of care in this setting, and thus extent of visceral KS is likely under-reported.^29^

In conclusion, a large fraction of patients with KS in this study had advanced stage disease at time of diagnosis, underscoring the need to target interventions to improve earlier diagnosis of KS. Additionally, within this group of advanced stage patients there is substantial heterogeneity, leading to questions about whether all patients with advanced KS require the same treatment strategy. Further studies are needed to confirm this clinical heterogeneity, and determine whether this heterogeneity is predictive of outcomes and should be included in a modified KS staging system.

## Data Availability

The data is stored on Dropbox.com and a de-identified version can be provided on request.

